# Distinct neural substrates of affective distress and communicative disability in head and neck cancer: a cross-sectional 18F-FDG PET study

**DOI:** 10.64898/2026.05.07.26352637

**Authors:** Antonis Tsionis, Efthymios Kyrodimos, Sofia Chatziioannou, Charalambos Papageorgiou

**Affiliations:** 1st Department of Psychiatry, Eginition Hospital, School of Medicine, National and Kapodistrian University of Athens, Vasilissis Sofias Ave. 72-74, Athens 11528, Greece; 1st Department of Otolaryngology Head and Neck Surgery, Hippokrateio Hospital, National and Kapodistrian University of Athens, Vasilissis Sofias Ave. 114, Athens 11527, Greece; Chair Department of Nuclear Medicine and Molecular Imaging, 2nd Department of Radiology, Medical School, National and Kapodistrian University of Athens, General University Hospital “ATTIKON”, 1 Rimini Street, Athens 12462, Greece; Director of PET/CT, Department of Nuclear Medicine, Biomedical Research Foundation of the Academy of Athens, 4 Soranou Ephessiou Street, Athens 11527, Greece; University Mental Health Neurosciences and Precision Medicine Research Institute “Costas Stefanis”, 4 Soranou Ephessiou Street, Athens 11527, Greece

**Keywords:** Head and neck cancer, Depression, FDG-PET, Brain metabolism, Broca, psychosomatic, voice

## Abstract

**Background:** Head and neck cancer (HNC) uniquely threatens communication through its impact on voice and speech. The neural systems linking depressive symptoms with perceived voice handicap remain poorly characterized. We examined whether these symptom domains show dissociable associations with regional brain glucose metabolism.

**Methods:** We conducted a cross-sectional ^18F^FDG-PET study in 63 HNC patients following diagnosis. Regional cerebral glucose metabolism (standardized uptake value ratios) was quantified in five a priori regions of interest: Broca’s area, Wernicke’s area, bilateral insula, and bilateral hippocampus. Depressive symptoms (Zung Self-Rating Depression Scale, SDS) and perceived voice handicap (Voice Handicap Index, VHI) were assessed. Spearman correlations with false discovery rate correction, partial correlations, and unique variance analyses were performed. Brain-behavior associations were examined using Spearman correlations with FDR correction.

**Results:** Depression and perceived voice handicap were strongly correlated (ρ = 0.64, p < 0.001) and exhibited partially dissociable metabolic correlates. Depressive symptom severity was associated with reduced metabolism in Broca’s area (ρ = −0.31, p_FDR = 0.041) and higher metabolism in the left insula (ρ = 0.36, p_FDR = 0.039), with graded insular elevation in moderate–severe depression (+14%, p = 0.008). Voice handicap was associated with reduced hippocampal metabolism (ρ = −0.34, p = 0.016 after covariate adjustment) in exploratory analyses.

**Conclusions:** Depression and voice handicap in HNC arise from partially independent neurobiological processes despite their clinical co-occurrence. This dissociation supports parallel rather than hierarchical supportive care pathways. Routine clinical imaging can be leveraged to generate testable hypotheses for psychosomatic and rehabilitation research.

Graphical abstract
Depressive symptoms were associated with reduced Broca’s area metabolism and elevated left insula metabolism, consistent with limbic-cortical dysregulation. Perceived voice handicap was uniquely associated with reduced bilateral hippocampal metabolism, implicating contextual memory and adaptation processes. Arrows indicate direction of association.

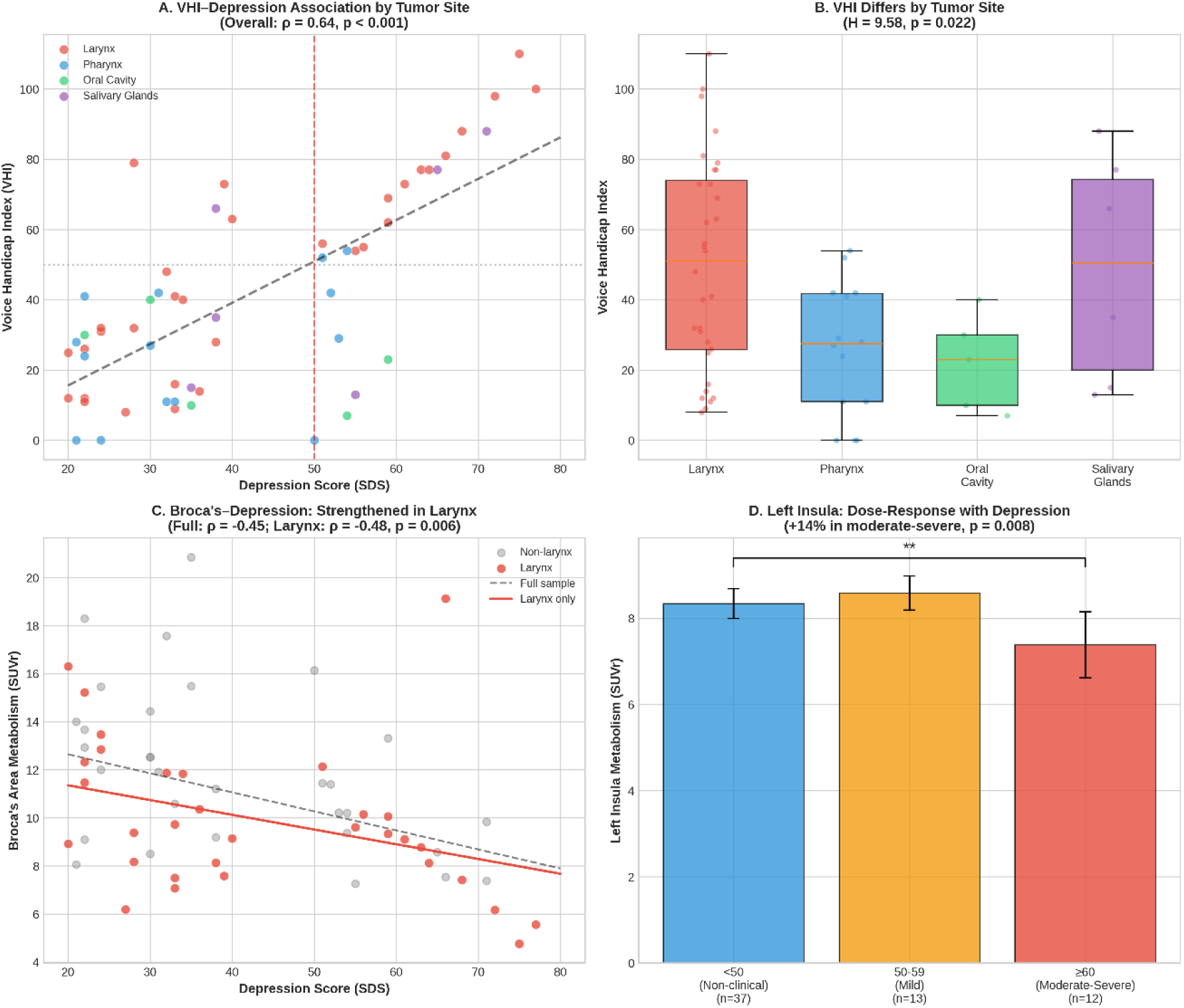

## 1. Introduction

Head and neck cancer (HNC) refers to a series of malignant tumors that occur above the clavicles, below the skull base, and in the anterior aspect of the neck vertebral column and treated categorically is the one of the most prevalent cancer worldwide, with 1,464,550 new cases and 487,993 deaths each year. [1,2]

HNC patients confront distinctive threats to core aspects of human identity and social functioning [3]. Voice impairment results from tumor location, disease progression, or effects of treatment on the larynx, pharynx, and surrounding structures, frequently disrupting communication capacity and contributing to psychological distress. Depression and perceived voice handicap are linked even at the time of diagnosis [4], and pretreatment communication ability independently predicts quality of life beyond tumor characteristics and depression severity [5].

Patients with HNC are approximately three times more likely to screen positive for depression compared with other cancer types [6]. A meta-analysis of 208 studies established that 19.5% report clinically significant depressive symptoms, with suicide incidence reaching 161 per 100,000 person-years [7]. The elevated screening rates do not translate into equivalent rates of diagnosed depression, as evidence demonstrates lower prevalence estimates when depression is assessed using diagnostic interviews compared with self-report instruments [8]. Pre-treatment depression or depressive symptoms in HNC predict worse overall survival (HR 1.33, 95% CI 1.16–1.52) [9].

Functional neuroimaging has advanced understanding of depression neurobiology. Recent ^18F^FDG-PET studies in mixed cancer populations have demonstrated correlations between regional brain glucose metabolism and depression severity [10], but remarkably little work has examined the neural correlates of depression or functional disability specifically in HNC patients. No study has examined whether depression and perceived voice handicap exhibit dissociable neural substrates. ^18F^FDG-PET/CT is already routinely performed in HNC patients as part of standard oncological staging and treatment planning, presenting an opportunity to extract neurobiological information from imaging acquired in routine care without additional burden to patients.

We selected regions of interest based on theoretical relevance. Broca’s area participates in prefrontal control systems and has shown depression-related dysfunction during verbal fluency paradigms [11,12]. Wernicke’s area supports speech comprehension and serves as a node of the language network, relevant to communication-related distress. [13,14] The insula plays a critical role in interoception and subjective emotional experience [15,16] and shows structural and functional abnormalities in depression [17,18]. The hippocampus is implicated in stress sensitivity, contextual processing, and depressive illness [19,20].

Using ^18F^FDG-PET imaging acquired during routine oncological staging, we examined regional brain metabolism in speech-related and affective processing regions. We hypothesized that voice handicap would correlate with depression severity, and examined whether they show overlapping or dissociable brain metabolic correlates. We predicted Broca’s area metabolism would associate with depression, Wernicke’s area with both domains, and hippocampal metabolism with both. Insula associations were explored without directional hypotheses.

## 2. Methods

### 2.1 Participants

Adults with histopathologically confirmed HNC were recruited from the A’ University Otorhinolaryngology Clinic, Hippokration General Hospital, Athens, in collaboration with the A’ University Psychiatric Clinic, Eginition Hospital. Eligibility required PET/CT imaging performed as part of routine oncological staging. Exclusion criteria were documented brain metastases, history of major neurological disorder, or pre-existing severe psychiatric ilness. The study was approved by the institutional ethics committees (ΨΓΟΖ46Ψ8Ν2-Α1Δ, Eginition) and all participants provided written informed consent,

### 2.2 Measures

#### 2.2.1 Primary Outcomes

Regional cerebral glucose metabolism was assessed using ^18F^F-fluorodeoxyglucose positron emission full body tomography (^18F^FDG-PET). Images were analyzed using RadiAnt DICOM Viewer, and standardized uptake values (SUVs) were calculated and normalized for body mass and whole-brain mean uptake, yielding SUV ratios (SUVr) for inter-individual variability in global metabolism. Regions of interest (ROIs) were selected a priori based on their theoretical relevance. A nuclear medicine specialist manually delineated ROIs. Prior to adopting this approach, automated segmentation was tested in a subset of 10 participants with comparable estimates. Given the equivalent results and greater anatomical precision and feasibility, manual delineation was used for the full sample. The following ROIs were examined: Broca’s area, Wernicke’s area, left and right insula and bilateral hippocampus.

Depressive symptoms were assessed using the Zung Self-Rating Depression Scale (SDS), a 20-item self-report questionnaire rated on a four-point Likert scale, total scores range from 20 to 80, with higher scores indicating greater symptom severity. Scores ≥50 indicated clinically significant depression, and scores ≥60 indicated moderate-to-severe depression. [21,22]

Perceived voice-related disability was assessed using the 30-item Voice Handicap Index (VHI). Items are rated on a five-point Likert scale, with total scores ranging from 0 to 120 and higher scores reflecting greater perceived voice handicap. [23]

#### 2.2.1 Secondary measures

##### General Psychological Distress

General psychological distress was assessed using the Symptom Checklist-90-Revised (SCL-90-R), a 90-item self-report measure rated on a five-point Likert scale. The instrument yields scores for nine symptom dimensions and a global severity index. [24,25].

### 2.3 Statistical Analysis

Data were explored descriptively to inform the selection of non-parametric correlation analyses. Associations (VHI and SDS) were examined using Spearman rank correlations. Ten tests were performed (5 ROIs × 2 outcomes), with Benjamini–Hochberg false discovery rate (FDR) correction applied to control for multiple comparisons. Bootstrap 95% confidence intervals were estimated using 2,000 resamples. Partial correlations adjusted for age and sex were computed to assess confounding.

Nested linear regression models were estimated for each ROI. A full model including VHI, SDS, age, and sex was compared with reduced models excluding either clinical predictor. Unique variance was quantified as the change in explained variance (ΔR²), reflecting the semi-partial contribution of each predictor beyond shared variance. Model residuals were examined to verify assumptions; given acceptable residual distributions, standard OLS regression was used.

Examining tumor site effects, tumor location was dichotomized as laryngeal versus non-laryngeal. Secondary analyses examined outcomes across the four main tumor sites separately. Tumor staging was recorded according to the TNM classification system. T stage was categorized as early (T1–T2) versus advanced (T3–T4), and nodal status was categorized as node-negative (N0) versus node-positive (N+). Associations were examined using Mann-Whitney U tests for dichotomous comparisons and Kruskal-Wallis tests for comparisons across the four main tumor sites. Chi-square tests examined differences in depression prevalence across tumor sites. Associations between TNM staging parameters and clinical outcomes were examined using Mann-Whitney U tests.

Participants were also grouped according to established SDS thresholds (<50, 50–59, ≥60). Differences in ROI metabolism were assessed using Kruskal–Wallis tests for three-group comparisons and Wilcoxon rank-sum tests for two-group contrasts, with post hoc Dunn’s tests and FDR correction where applicable.

Sensitivity analyses examined statistical outliers (|z| > 3), sex-by-predictor interaction effects, non-linear associations (quadratic terms). Secondary analyses examined replication of primary findings using alternative depression-related measures (SCL-90-R Depression subscale and EPQ Neuroticism); given the reduced sample size (n = 57–60) and exploratory nature, these were not FDR-corrected. To assess whether primary brain-behavior associations were confounded by tumor site, correlations were examined within the laryngeal cancer subgroup separately.

Effect sizes for correlations were interpreted using Gignac and Szodorai (2016) guidelines for individual differences research: small |ρ| ≥ 0.10, medium |ρ| ≥ 0.20, large |ρ| ≥ 0.30.[26]. Tests were two-tailed with α = 0.05.

All analyses were conducted in R (version 4.5.1).

## 3. Results

### 3.1 Sample Characteristics

The sample consisted of 63 head and neck cancer patients, 39 males (61.9%), with a mean age of 61.6 ± 11.2 years (range: 35–81). Mean VHI score was 41.0 ± 28.6 (range: 0–110), and mean SDS score was 41.6 ± 17.4 (range: 20–77). Clinically significant depressive symptoms (SDS ≥50) were present in 25 patients (39.7%), including 12 patients (19.0%) meeting criteria for moderate-to-severe depression (SDS ≥60).

Regarding tumor site, 32 patients (52.5%) had laryngeal cancer, 14 (23.0%) pharyngeal cancer, 6 (9.8%) salivary gland tumors, 5 (8.2%) oral cavity tumors, and 4 (6.5%) other or unknown primary sites. Among 52 patients with available TNM staging, 28 (53.8%) had early-stage tumors (T1–T2) and 24 (46.2%) had advanced-stage tumors (T3–T4). Nodal involvement was present in 21 of 50 patients with available N staging (42.0%). (Table 1)

**Table 1.** Sample characteristics.

| Variable | n | Mean (SD) or n (%) | Range |
| --- | --- | --- | --- |
| **Demographics** |  |  |  |
| Age (years) | 63 | 61.6 (11.2) | 35-81 |
| Sex, Male | 39 | (61.9%) |  |
| Sex, Female | 24 | (38.1%) |  |
| **Tumor Characteristics** |  |  |  |
| Primary Site |  |  |  |
| Larynx | 32 | (52.5%) |  |
| Pharynx | 14 | (23%) |  |
| Salivary Glands | 6 | (9.8%) |  |
| Oral Cavity | 5 | (8.2%) |  |
| Other/Unknown | 4 | (6.5%) |  |
| T Stage (n=52) |  |  |  |
| T1-T2 (Early) | 28 | (53.8%) |  |
| T3-T4 (Advanced) | 24 | (46.2%) |  |
| N Stage (n=50) |  |  |  |
| N0 | 29 | (58%) |  |
| N+ | 21 | (42%) |  |
| **Clinical Outcomes** |  |  |  |
| Voice Handicap Index (VHI) | 63 | 41.0 (28.6) | 0 - 110 |
| Zung Self-Rating Depression Scale | 63 | 41.6 (17.4) | 20 - 77 |
| Clinical threshold($\geq 50$) | 25 | (39.7%) | |
| Modere-severe ($\geq 60$) | 12 | (19%) | |

### 3.2 Association Between Voice Handicap Index and Depression Symptoms

Voice handicap and depressive symptom severity were strongly correlated (ρ = 0.64, p < 0.001), corresponding to approximately 41% shared variance. This association remained unchanged after adjustment for age and sex (ρ partial = 0.64, p < 0.001). (Figure 1)

**Figure 1.**
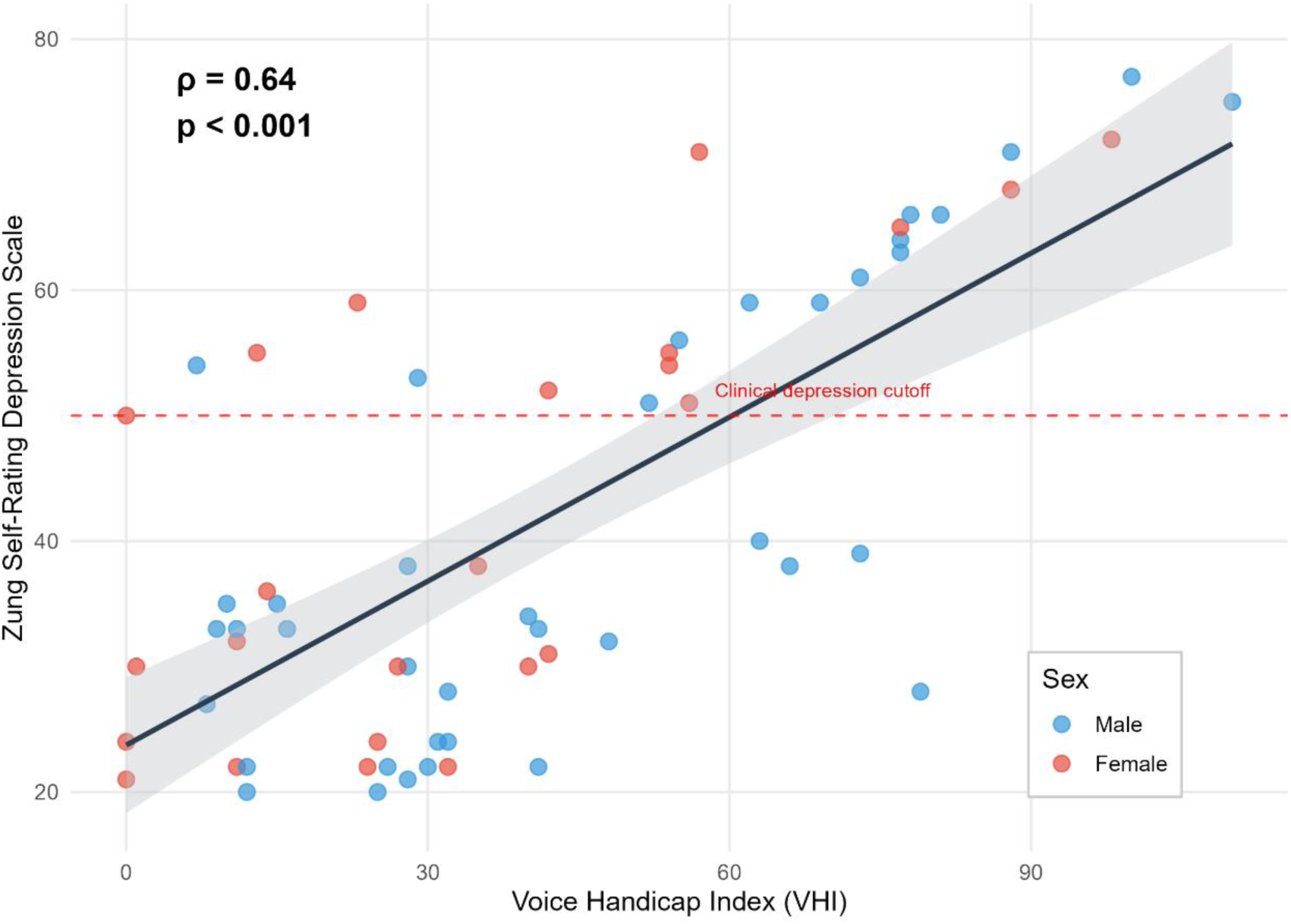
Association Between Voice Handicap and Depression. The solid line denotes the fitted linear trend, with shaded bands indicating the 95% confidence interval. The dashed horizontal line marks the clinical cutoff for depressive symptoms (SDS = 50). The strength of association is quantified using Spearman’s rank correlation (ρ = 0.64, p < 0.001).

### 3.3 Tumor Characteristics and Clinical Outcomes

Perceived voice handicap differed significantly by tumor site. In the primary dichotomous analysis, patients with laryngeal cancer reported substantially higher VHI scores compared to patients with non-laryngeal tumors (50.0 ± 30.1 vs. 29.6 ± 23.4; Mann-Whitney U = 603.5, p = 0.009, Cohen’s d = 0.75). Secondary analyses across the four main tumor sites confirmed this pattern (Kruskal-Wallis H = 9.58, p = 0.022), with post-hoc comparisons showing laryngeal cancer patients reporting significantly higher voice handicap than pharyngeal (p = 0.011, d = 0.89) and oral cavity patients (p = 0.037, d = 0.98). Notably, salivary gland patients showed similarly elevated VHI scores to laryngeal patients (49.0 vs. 50.0; p = 0.94), though this subgroup was small (n = 6). (Figure 2A; Table 2)

**Figure 2.**
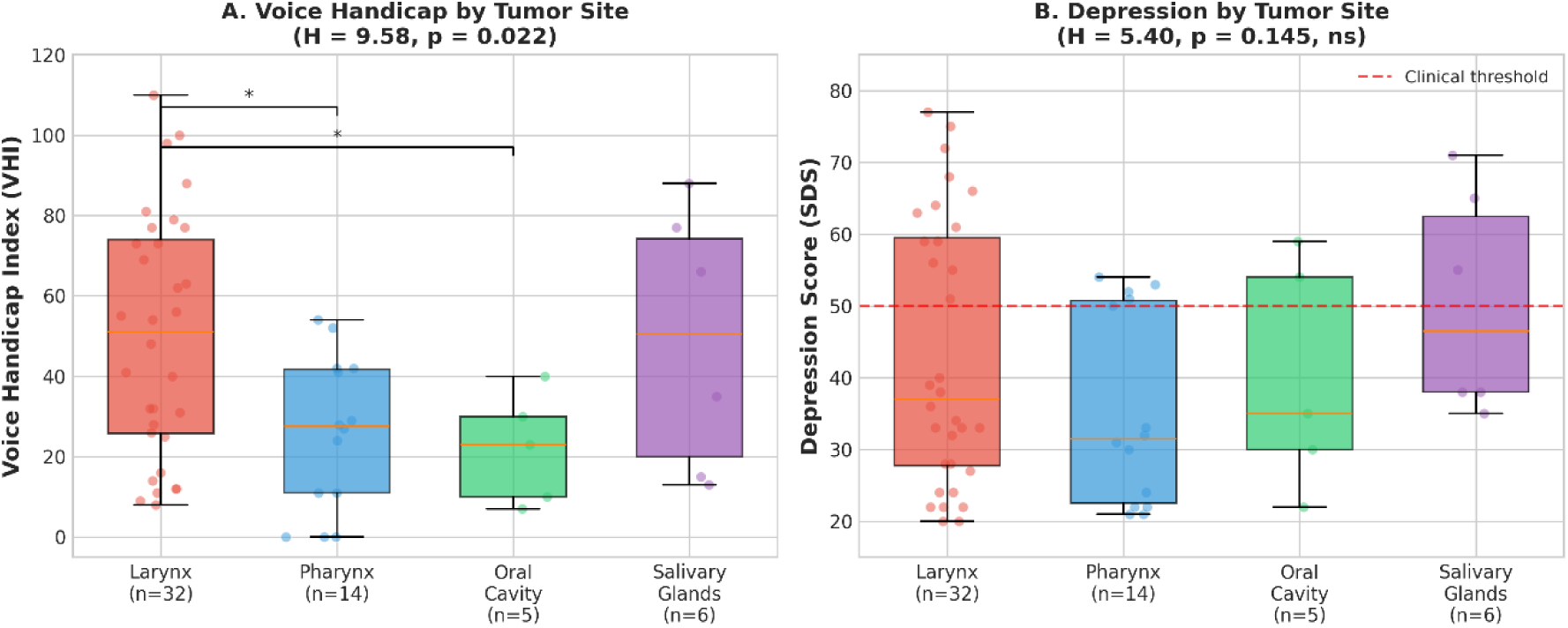
Depression and voice handicap by tumor site. Boxplots of clinical outcomes by tumor site. (A) Voice Handicap Index scores across four primary tumor sites. Laryngeal cancer patients reported significantly higher voice handicap than pharyngeal (p = 0.011) and oral cavity (p = 0.037) patients. (B) Zung Self-Rating Depression Scale scores across the same tumor sites. Depression severity did not differ significantly by tumor site (p = 0.145).

**Table 2.** Clinical outcomes by tumor characteristics.

| Variable | Group | n | VHI Mean (SD) | p | SDS Mean (SD) | p |
| --- | --- | --- | --- | --- | --- | --- |
| Tumor Site | Larynx | 32 | 50.0 (30.1) | **0.009** | 43.2 (18.5) | 0.333 |
|  | Non-larynx | 27 | 29.6 (23.4) |  | 38.9 (15.1) |  |
| T Stage | Early (T1–T2) | 28 | 36.5 (25.7) | 0.168 | 40.6 (17.2) | 0.228 |
|  | Advanced (T3–T4) | 24 | 49.2 (32.3) |  | 46.5 (18.8) |  |
| N Stage | N0 | 29 | 51.0 (30.9) | 0.016* | 43.4 (19.4) | 0.701 |
|  | N+ | 21 | 29.6 (22.6) |  | 42.6 (16.7) |  |

**Note.** VHI: Voice Handicap Index; SDS: Zung Self-Rating Depression Scale. P values from Mann-Whitney U tests.

In contrast, depressive symptom severity did not differ by tumor site in either dichotomous (Larynx: 43.2 ± 18.5 vs. Other: 38.9 ± 15.1; U = 496.0, p = 0.333) or four-group analyses (H = 5.40, p = 0.145). Rates of clinically significant depression were comparable across tumor sites: larynx 40.6%, pharynx 35.7%, oral cavity 40.0%, and salivary glands 50.0% (χ² = 0.08, p = 0.99). (Figure 2B; Table 2)

Tumor staging. Neither T stage nor N stage was significantly associated with depressive symptom severity. Advanced T stage (T3–T4) showed a trend toward higher depression scores compared to early-stage tumors (T1–T2), but this did not reach significance (46.5 ± 18.8 vs. 40.6 ± 17.2; U = 270.0, p = 0.228). Nodal involvement was also not associated with depression (N0: 43.4 ± 19.4 vs. N+: 42.6 ± 16.7; p = 0.701).

VHI scores did not differ significantly by T stage (Early: 36.5 ± 25.7 vs. Advanced: 49.2 ± 32.3; U = 260.5, p = 0.168). An apparent association between N stage and VHI (N0: 51.0 ± 30.9 vs. N+: 29.6 ± 22.6; U = 427.0, p = 0.016) was confounded by differential distribution of tumor sites: laryngeal cancers were predominantly N0 (85.2%), whereas non-laryngeal tumors were predominantly node-positive (72.7%; χ² = 14.52, p < 0.001). When stratified by tumor site, N stage was not associated with VHI within either subgroup.

### 3.4 Regional Brain Metabolism and Primary Clinical Outcomes

Regional brain metabolism showed differential associations with SDS and VHI. (Table 3 and Figure 3)

**Figure 3.**
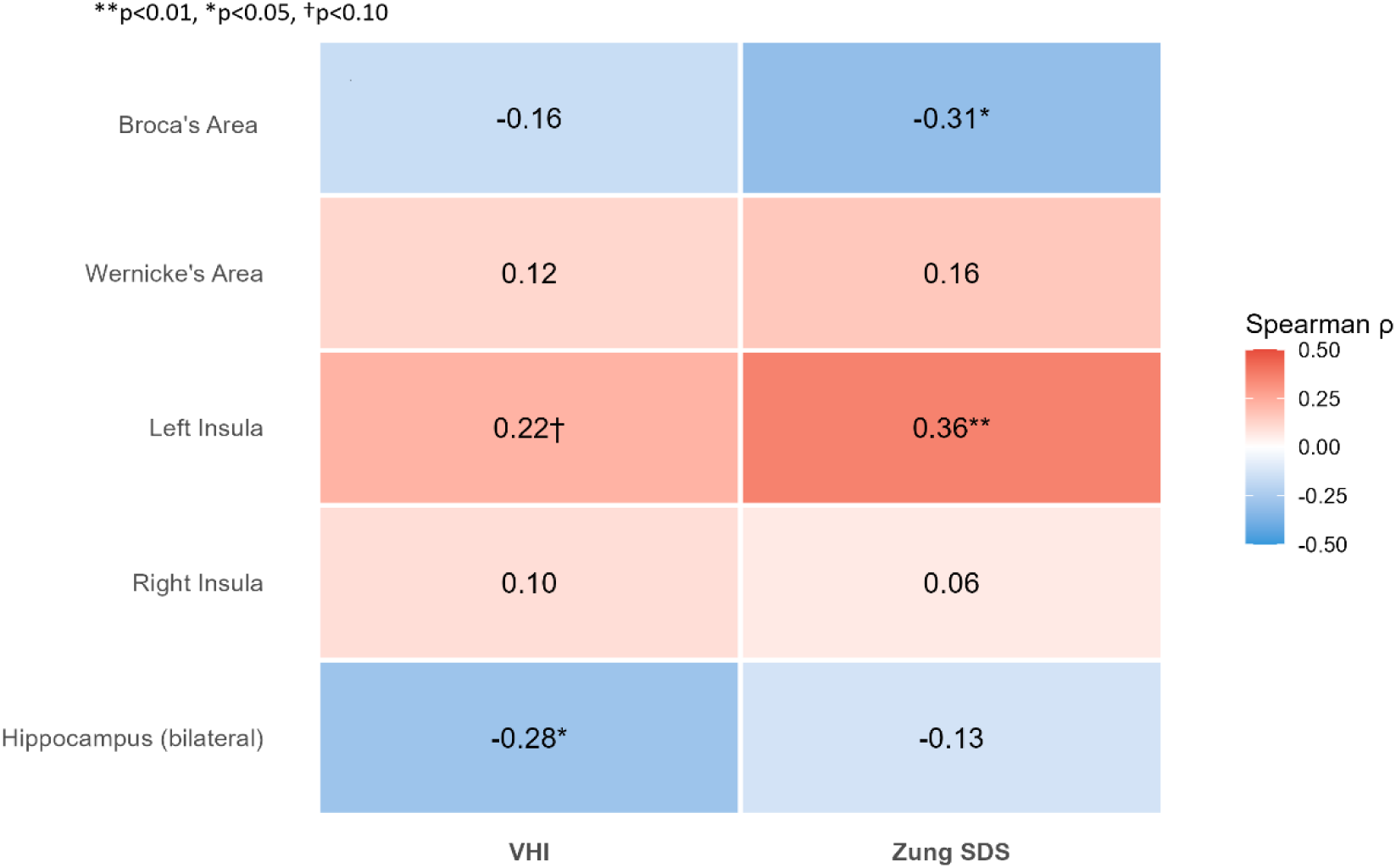
Cells display Spearman’s rank correlation coefficients (ρ) between regional standardized uptake value ratios (SUVr; normalized to whole-brain mean) and clinical measures. Values are color-coded by effect size (red = positive, blue = negative). Significance thresholds are denoted as **\*\***p < 0.01, *p < 0.05, and †p < 0.10.

**Table 3.** Correlations Between Local Brain Metabolism and Clinical Outcomes.

| ROI | Outcome | n | $\rho$ | 95% CI | p | p(FDR) | Significance | Effect |
| --- | --- | --- | --- | --- | --- | --- | --- | --- |
| Broca's Area | Zung SDS | 63 | -0.307 | [-0.52, -0.05] | 0.014 | 0.041 | * | Medium |
| Left Insula | Zung SDS | 62 | 0.362 | [0.11, 0.57] | 0.004 | 0.039 | ** | Medium |
| Hippocampus (bilateral) | VHI | 53 | -0.281 | [-0.52, -0.00] | 0.041 | 0.137 | † | Small |
| Wernicke's Area | Zung SDS | 63 | 0.162 | [-0.10, 0.40] | 0.204 | 0.345 |  | Small |
| Hippocampus (bilateral) | Zung SDS | 53 | -0.130 | [-0.40, 0.15] | 0.354 | 0.449 |  | Small |
| Right Insula | Zung SDS | 62 | 0.063 | [-0.19, 0.31] | 0.629 | 0.629 |  | Negligible |
| Broca's Area | VHI | 63 | -0.161 | [-0.43, 0.10] | 0.207 | 0.345 |  | Small |
| Left Insula | VHI | 62 | 0.221 | [-0.02, 0.44] | 0.084 | 0.210 |  | Small |
| Wernicke's Area | VHI | 63 | 0.118 | [-0.14, 0.38] | 0.359 | 0.449 |  | Small |
| Right Insula | VHI | 62 | 0.096 | [-0.17, 0.34] | 0.458 | 0.509 |  | Negligible |

**Note.** Values are Spearman's rank correlation coefficients ( $\rho$ ) between regional brain metabolism (SUVr, normalized to whole-brain mean) and clinical outcomes. Ninety-five percent confidence intervals were estimated using 2,000 bootstrap resamples (percentile method). P values were adjusted for multiple comparisons across the 10 a priori tests (5 ROIs × 2 outcomes) using the Benjamini–Hochberg false discovery rate (FDR) procedure. Significance is denoted as **\*\*** $p < 0.01$ , **\*** $p < 0.05$ , and **†** $p < 0.10$ . Effect size labels follow conventional thresholds for Spearman correlations (small $\geq 0.10$ , medium $\geq 0.30$ , large $\geq 0.50$ )

Broca’s area metabolism showed a negative correlation with SDS scores (ρ = −0.31, 95% CI [−0.42, −0.15], p = 0.014), survived FDR correction (p FDR = 0.041). The association remained after adjustment for age and sex (ρ partial = −0.31, p = 0.016). (Table 4)

**Table 4.** Partial correlations (Spearman’s ρ), adjusted for age and sex (FDR-corrected).

| ROI | Outcome | n | $\rho$ (partial) | p | p(FDR) | Significance |
| --- | --- | --- | --- | --- | --- | --- |
| Broca's Area | Zung SDS | 63 | -0.308 | 0.016 | 0.041 | * |
| Left Insula | Zung SDS | 62 | 0.365 | **0.004** | 0.041 | ** |
| Hippocampus (bilateral) | VHI | 53 | -0.337 | 0.016 | 0.043 | * |
| Left Insula | VHI | 62 | 0.228 | 0.080 | 0.201 |  |
| Broca's Area | VHI | 63 | -0.185 | 0.155 | 0.292 |  |
| Wernicke's Area | Zung SDS | 63 | 0.176 | 0.175 | 0.292 |  |
| Wernicke's Area | VHI | 63 | 0.148 | 0.254 | 0.363 |  |
| Hippocampus (bilateral) | Zung SDS | 53 | -0.124 | 0.384 | 0.480 |  |
| Right Insula | VHI | 62 | 0.087 | 0.507 | 0.563 |  |
| Right Insula | Zung SDS | 62 | 0.069 | 0.601 | 0.601 |  |

**Note.** Values are Spearman's partial correlation coefficients ( $\rho$ ), adjusted for age and sex. P values were adjusted for multiple comparisons across the 10 a priori tests (5 ROIs × 2 outcomes) using the Benjamini–Hochberg false discovery rate (FDR) procedure. Significance is denoted as \*\* $p < 0.01$ and $p < 0.05$ . Only associations surviving FDR correction are marked as significant.

Left insula metabolism showed a positive association with depressive symptom severity (ρ = 0.36, 95% CI [0.11, 0.57], p = 0.004), which survived FDR correction (p_FDR = 0.039) and remained significant after adjustment for age and sex (ρ_partial = 0.37, p = 0.004)

Bilateral hippocampal metabolism showed a negative association with VHI (ρ = −0.28, 95% CI [−0.52, −0.01], p = 0.041), which did not survive FDR correction (p_FDR = 0.137). After adjustment for age and sex, this association strengthened and reached significance (ρ partial = −0.34, p = 0.016). (Table 4)

Metabolism in Wernicke’s area and the right insula was not significantly correlated with either SDS or VHI scores (all p_FDR > 0.20).

### 3.5 Unique Variance Analysis

For Broca’s area, SDS explained 6.6% of unique variance (ΔR² = 6.6%), whereas VHI explained 0.2% (ΔR² = 0.2%). For the left insula, SDS explained 9.3% of unique variance (ΔR² = 9.3%), whereas VHI explained 0.8% (ΔR² = 0.8%). For the bilateral hippocampus, VHI explained 6.2% of unique variance (ΔR² = 6.2%), whereas SDS explained 0.3% (ΔR² = 0.3%). (Table 5)

**Table 5.**
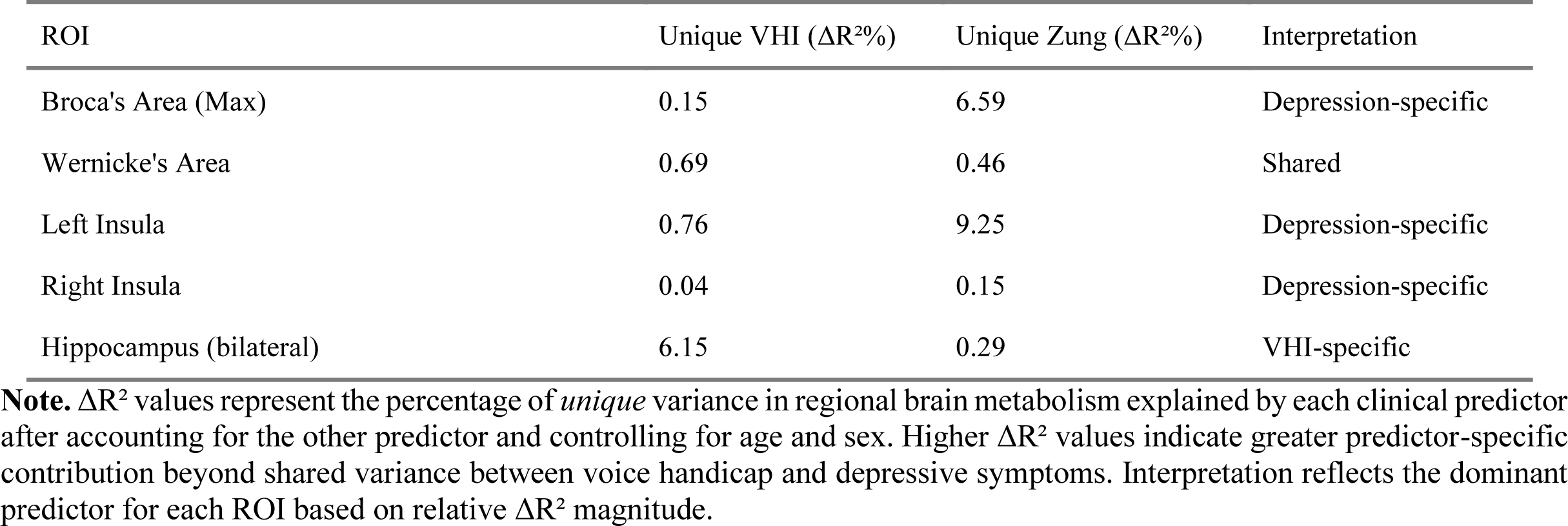
Unique variance analysis (ΔR²%).

### 3.6 Regional Brain Metabolism by Tumor Site

Regional brain metabolism also differed by tumor site. Broca’s area metabolism was significantly lower in laryngeal cancer patients compared to other sites (H = 9.97, p = 0.019), with post-hoc comparisons showing lower metabolism than pharyngeal (9.93 vs. 12.53; p = 0.013) and oral cavity patients (9.93 vs. 12.87; p = 0.024). Similar patterns were observed for Wernicke’s area (H = 9.29, p = 0.026) and right insula (H = 9.19, p = 0.027). Left insula showed a trend (H = 7.25, p = 0.065). Hippocampal metabolism did not differ by tumor site (p > 0.28). (Table 7)

### 3.7 Depression Severity Group Comparisons

Participants were categorized according to SDS thresholds: non-clinical (<50), mild depression (50–59), and moderate-to-severe depression (≥60). Groups did not differ in age (p = 0.26) or sex distribution (p = 0.60).

VHI scores differed across depression severity groups (Kruskal–Wallis H = 28.35, p < 0.001). Mean VHI scores were 28.1 ± 19.4 in the non-clinical group, 39.7 ± 22.7 in the mild depression group, and 83.7 ± 14.1 in the moderate-to-severe depression group.

Left insula metabolism differed across depression severity groups (Kruskal–Wallis H = 7.89, p = 0.019). Mean SUVr values were 1.14 in the non-clinical group, 1.18 in the mild depression group, and 1.30 in the moderate-to-severe depression group. Post-hoc comparisons showed a significant difference between the moderate-to-severe and non-clinical groups (p = 0.008). In binary comparison, moderate-to-severe depression (≥60) versus all others showed significantly higher left insula metabolism (p = 0.008, p_FDR = 0.041).

Broca’s area metabolism did not differ across depression severity groups (Kruskal–Wallis H = 3.08, p = 0.214).

Hippocampal metabolism did not differ across depression severity groups (Kruskal–Wallis H = 1.33, p = 0.515). (Table 6)

**Table 6.** ROI Metabolism: Moderate-to-severe depression symptoms (≥60) vs. lower symptoms.

| ROI | SDS < 60 | SDS $\geq 60$ | Diff (%) | p | p FDR |
| --- | --- | --- | --- | --- | --- |
| Left Insula | 1.15<br>(0.17) | 1.30 (0.22) | +12.5% | 0.008 | 0.041* |
| Wernicke's Area | 1.25<br>(0.16) | 1.35 (0.19) | +8.6% | 0.026 | 0.064 |
| Broca's Area (Max) | 1.58<br>(0.38) | 1.48 (0.30) | -6.5% | 0.298 | 0.306 |
| Right Insula | 1.14<br>(0.12) | 1.19 (0.17) | +3.9% | 0.306 | 0.306 |
| Hippocampus | 0.79<br>(0.11) | 0.70 (0.09) | -10.5% | 0.299 | 0.306 |

**Note.** Values are mean (SD) standardized uptake value ratios (SUVr), normalized to whole-brain mean uptake. Participants dichotomized into moderate-to-severe depression (Zung Self-Rating Depression Scale $\geq 60$ ) versus all others ( $< 60$ ). Group differences were assessed using Wilcoxon rank-sum tests. p values were adjusted for multiple comparisons across regions of interest using the Benjamini–Hochberg false discovery rate (FDR) \*p FDR < 0.05.

### 3.8 Sex and Sensitivity Analyses

After exclusion of statistical outliers (|z| > 3), the correlations between regional metabolism and SDS remained: Broca’s area (ρ = −0.25, p = 0.047) and left insula (ρ = 0.35, p = 0.006). No sex-by-predictor interaction terms reached statistical significance (all p > 0.32). Inclusion of quadratic terms did not improve model fit.

To assess whether primary brain-clinical associations were confounded by tumor site, sensitivity analyses were conducted within the laryngeal cancer subgroup (n = 32). The correlation between VHI and depression was notably stronger within laryngeal cancer patients (ρ = 0.83, p < 0.001) compared to the full sample (ρ = 0.64). The association between Broca’s area metabolism and depression was also strengthened (ρ = −0.48, p = 0.006), suggesting this relationship is particularly robust in patients with tumors directly affecting the larynx. Right hippocampal metabolism showed a significant negative correlation with depression within this subgroup (ρ = −0.41, p = 0.033). (Table 7)

**Table 7.** Associations Between Regional Brain Metabolism and Clinical Measures Within Each Tumor Site.

| ROI | Larynx<br>(n=32) |  | Pharynx<br>(n=14) |  | Oral Cavity<br>(n=5) |  | Salivary<br>(n=6) |  |
| --- | --- | --- | --- | --- | --- | --- | --- | --- |
|  | VHI | SDS | VHI | SDS | VHI | SDS | VHI | SDS |
| Broca's Area | -0.39* | -0.48** | -0.32 | -0.16 | 0.10 | -0.30 | -0.09 | -0.81* |
| Wernicke's Area | -0.13 | -0.19 | -0.32 | -0.06 | 0.50 | 0.20 | -0.37 | -0.93** |
| Left Insula | -0.22 | -0.16 | -0.33 | 0.17 | 0.80 | -0.60 | -0.71 | -0.90* |
| Right Insula | -0.19 | -0.29 | -0.37 | -0.11 | -0.10 | -0.20 | -0.37 | -0.70 |
| L Hippocampus | -0.32 | -0.27 | -0.68* | -0.20 | -0.90* | 0.50 | -0.30 | -0.98** |
| R Hippocampus | -0.34 | -0.41* | -0.54 | -0.31 | -0.70 | 0.60 | -0.70 | -0.82 |
| **VHI ~ SDS** | **$\rho = 0.83^*$** |  | **$\rho = 0.47$** |  | **$\rho = -0.60$** |  | **$\rho = 0.64$** |  |

**Note.** Values are Spearman's correlation coefficients ( $\rho$ ) between regional brain metabolism and clinical measures within each tumor site; VHI= Voice Handicap Index; SDS= Zung Self-Rating Depression Scale. Subgroup sizes are small; correlations are descriptive and should be interpreted cautiously. \* $p < 0.05$ , \*\* $p < 0.01$ , \*\*\* $p < 0.001$ .

Sensitivity analyses using the SCL-90-R depression subscale confirmed that depression scores did not differ by tumor site (Larynx vs. Other: p = 0.89; four-group comparison: p = 0.77).

### 3.9 Secondary Psychometric Analyses

Secondary psychometric measures were available for 47–50 participants depending on the scale. Owing to the reduced sample and multiple comparisons, analyses were considered exploratory and no findings survived FDR correction.

For Broca’s area, correlations with the SCL-90-R depression subscale were negative and small (ρ = −0.18), with no association with SCL-90-R anxiety.

Left insula metabolism showed a positive but non-significant correlation with SCL-90-R depression (ρ = 0.15, p = 0.058) and no association with anxiety (ρ = 0.02, p = 0.91).

Hippocampus showed negative correlations with several SCL-90-R subscales, including psychoticism (ρ = −0.53, p = 0.008) and interpersonal sensitivity (ρ = −0.46, p = 0.023), though these did not survive multiple-comparison correction.

SCL-90-R subscales correlated with SDS scores (ρ = 0.53–0.75, all p < 0.01). Associations with VHI were smaller, with only the SCL-90-R depression subscale reaching nominal significance (ρ = 0.41, p = 0.043).

## 4. Discussion

This study provides the first neuroimaging evidence that depression and perceived voice handicap in HNC, though strongly correlated behaviorally, show partially dissociable brain metabolic correlates. Three principal findings emerged: voice handicap was strongly associated with tumor site, with laryngeal cancer patients reporting substantially higher perceived disability than patients with pharyngeal or oral cavity tumors, a large effect (d = 0.75–0.98) reflecting the direct anatomical impact on the phonatory organ. Second, depressive symptom severity was independent of tumor site, with comparable rates of clinically significant depression across all tumor locations. Third, depression and voice handicap showed dissociable associations with regional brain metabolism, suggesting that affective distress and perceived functional disability rely on overlapping but non-identical neural networks.

Broca’s area metabolism showed a negative association with depressive symptom severity and specific to depression rather than voice handicap. This pattern aligns with the hypofrontality hypothesis of depression, which posits reduced frontal metabolic activity as a core feature of the disorder [27,28]. PET studies report reduced prefrontal glucose metabolism in depression [29,30], with meta-analyses confirming left frontal hypofunction [30,31].

Extending hypo-frontality to speech production networks is theoretically coherent given that depression is associated with psychomotor retardation, reduced verbal fluency, and poor spontaneous speech [32]. These are further supported by systematic reviews demonstrating reduced left inferior frontal gyrus activation during verbal fluency in depression [12,33], with recent machine learning studies identifying Broca’s area as a key discriminative feature for major depressive disorder classification [34]. Moreover, the frequent co-occurrence of aphasia and depression following left frontal lesions supports the plausibility that integrity of left frontal speech-related systems is clinically relevant to mood [35].

Notably, laryngeal cancer patients showed lower regional brain metabolism in Broca’s area, Wernicke’s area, and bilateral insula compared to pharyngeal and oral cavity patients. This pattern could reflect reduced engagement of speech-related circuitry due to voice impairment, chronic stress effects secondary to communication difficulties, or pre-existing neural characteristics. The strengthening of brain-behavior correlations within the laryngeal subgroup supports the hypothesis that fronto-speech circuitry is especially relevant for understanding depression in patients whose primary tumor directly affects voice production.

In HNC, depressed patients may show reduced motivation to communicate, lower engagement with compensatory strategies, and social withdrawal, all of which could reduce use-dependent activation of motor-speech circuitry. Alternatively, lower baseline left frontal function may represent a shared vulnerability that both increases susceptibility to depressive symptoms and reduces adaptive capacity in the face of communication changes.

Left insula metabolism showed a positive association specific to depression rather than voice handicap. A graded severity-dependent increase relationship strengthened interpretability: with the moderate-to-severe group showing significantly higher metabolism. This is consistent with evidence that insula metabolism may serve as a treatment-response biomarker, with elevated baseline insula activity predicting better response to cognitive behavioral therapy versus medication [36].

This finding is consistent with evidence implicating the insula in interoceptive processing and subjective emotional experience [15,16]. Structural meta-analyses report insular gray matter abnormalities in major depressive disorder [18], and functional studies demonstrate altered insula responses during emotional and interoceptive tasks as well as resting-state conditions [17,37].

In the present HNC cohort, elevated left insula metabolism may reflect heightened interoceptive monitoring, negative affective processing, or rumination-like mechanisms in depression. The opposite directionality observed for Broca’s area (hypometabolism) versus insula (hypermetabolism) is compatible with models proposing frontal hypoactivity coupled with limbic/paralimbic hyperactivity in depression [28,38]. This “limbic-cortical dysregulation” model has received support from treatment studies showing normalization of this imbalance with successful antidepressant intervention [39].

Right insula metabolism was not significantly associated with depression or voice. The lateralization to left insula may be consistent with reports suggesting left-hemisphere predominance for aspects of depression-related interoceptive or verbally mediated emotional processing, though the broader literature is mixed [15].

Wernicke’s area metabolism did not show significant associations with depression or voice handicap. This is theoretically coherent because Wernicke’s area supports speech comprehension, phonological processing, and lexical-semantic retrieval [14,40], and comprehension systems may be less directly impacted by the vocal production impairment characteristic of HNC. A trend toward higher metabolism in moderate–severe depression was observed, but this requires replication.

Bilateral hippocampus metabolism showed a negative correlation with voice handicap. Unique variance analysis indicated specificity: voice handicap uniquely explained 6.2% of hippocampal metabolism variance, whereas depression explained only 0.3%.

This dissociation is noteworthy given the classical association of hippocampal alterations with depression [41,42]. In the present sample, hippocampal metabolism instead tracked perceived functional disability, suggesting that memory or contextual processing systems may contribute to how patients appraise and integrate voice-related limitations. Mechanistically, reduced hippocampal function could impair contextual discrimination, promoting overgeneralized disability perception across situations [43].

The hippocampus is highly susceptible to stress-induced changes, with chronic stress producing structural and functional alterations through glucocorticoid-mediated mechanisms [44,45]. Chronic stress exposure is associated with reduced hippocampal neurogenesis, dendritic remodeling, and impaired synaptic plasticity, processes that may contribute to the memory difficulties and contextual processing deficits observed in depression and chronic illness adaptation [46].

The hippocampus also contributes to autobiographical memory and self-related integration over time [47,48]. Difficulty integrating a “changed voice” into self-concept could increase perceived handicap. Furthermore, adaptation to voice change requires learning and consolidation of compensatory strategies, which may be reduced with lower hippocampal function.

The independence of depression from tumor site, with rates of 35–50% across all locations, suggests that psychological distress in HNC reflects common factors such as the cancer diagnosis itself, treatment burden, uncertainty, and social disruption.

Several aspects of the study strengthen confidence in the findings. Region-of-interest selection was theory driven, focusing on speech-related regions and systems implicated in affective processing and adaptation, rather than relying on broad exploratory voxel-wise approaches. This strategy reduced the multiple-testing burden. Correction for multiple comparisons using false discovery rate procedures supports the primary depression-related results. In addition, the use of several complementary analytic approaches, including partial correlations and unique variance analyses, allowed an interpretation of overlapping symptom domains.

Several limitations should be considered. The cross-sectional design does not allow conclusions regarding causality or ordering between brain metabolism and clinical symptoms. Sample size was modest, particularly for analyses involving smaller tumor site subgroups (oral cavity n = 5, salivary glands n = 6), and findings in these subgroups require replication. Although we demonstrated important associations between tumor site and voice handicap, the sample was insufficiently powered to fully disentangle tumor-specific effects from treatment effects, as treatment modality varied by tumor characteristics. Moreover, resting-state ^18F^FDG-PET reflects baseline metabolic activity and cannot capture task-specific neural engagement during speech production or emotional processing.

In summary, in HNC, depression and perceived voice handicap are closely related at the behavioral level but show partially dissociable metabolic correlates. Depressive symptoms were associated with increased left insula metabolism in a graded manner, alongside an association with reduced Broca’s area metabolism. In contrast, perceived voice handicap was specifically associated with reduced hippocampal metabolism, pointing to the involvement of contextual memory and adaptation processes. Together, these findings support the view that affective distress and perceived functional disability represent related but separable neurobiological dimensions in head and neck cancer.

From a translational perspective, ^18F^FDG-PET/CT is already integrated into standard HNC care pathways for oncological staging and treatment planning. This presents an opportunity for incidental assessment of brain metabolic markers associated with psychological distress without requiring additional imaging procedures. If replicated in longitudinal analyses, the observed metabolic patterns could potentially inform early identification of patients at elevated risk for depression or voice-related disability, enabling timely referral for psychological support or speech rehabilitation.

Clinically, this partial dissociation suggests that similar presentations of distress may arise from different underlying processes. Patients whose symptoms are dominated by affective and interoceptive distress may benefit most from targeted depression treatments, whereas patients whose primary burden is perceived voice handicap may require interventions emphasizing rehabilitation, contextual learning, and psychological adaptation to communication changes. In many cases, a combined approach will be necessary, but the relative emphasis may differ across individuals.

Future phases include a longitudinal study to examine whether baseline metabolic patterns predict subsequent changes in depression or perceived voice handicap over the course of treatment, and whether symptom improvement or worsening is accompanied by corresponding changes in regional metabolism. Future research could utilize task-based imaging during speech production and affective or interoceptive processing to help clarify the functional significance of the observed associations.

## CRediT author statement

Antonis Tsionis: Conceptualization, Methodology, Data collection, Formal analysis, Writing – Original Draft. Efthymios Kyrodimos: Supervision, Resources, Writing – Review & Editing. Sophia N. Chatziioannou: Investigation (^18F^FDG-PET imaging), Methodology, Writing – Review & Editing. Charalambos C. Papageorgiou: Supervision, Conceptualization, Writing – Review & Editing.

## Data Availability

All data produced in the present study are available upon reasonable request to the authors

## Data statement

The data used for this study are available upon request.

## References

[1] F. Bray, M. Laversanne, H. Sung, J. Ferlay, R.L. Siegel, I. Soerjomataram, A. Jemal, Global cancer statistics 2022: GLOBOCAN estimates of incidence and mortality worldwide for 36 cancers in 185 countries, CA. Cancer J. Clin. 74 (2024) 229–263. 10.3322/caac.21834.

[2] T. Zhou, W. Huang, X. Wang, J. Zhang, E. Zhou, Y. Tu, J. Zou, K. Su, H. Yi, S. Yin, Global burden of head and neck cancers from 1990 to 2019, iScience 27 (2024). 10.1016/j.isci.2024.109282.

[3] L.H.A. Korsten, F. Jansen, B.J.F. De Haan, D. Sent, P. Cuijpers, C.R. Leemans, I.M. Verdonck-de Leeuw, Factors associated with depression over time in head and neck cancer patients: A systematic review, Psychooncology. 28 (2019) 1159–1183. 10.1002/pon.5058.

[4] J.Y.K. Chan, L.L. Lua, H.H. Starmer, D.Q. Sun, E.S. Rosenblatt, C.G. Gourin, The relationship between depressive symptoms and initial quality of life and function in head and neck cancer, The Laryngoscope 121 (2011) 1212–1218. 10.1002/lary.21788.

[5] C. Sauder, M. Kapsner-Smith, C. Baylor, K. Yorkston, N. Futran, T. Eadie, Communicative Participation and Quality of Life in Pretreatment Oral and Oropharyngeal Head and Neck Cancer, Otolaryngol. Neck Surg. 164 (2021) 616–623. 10.1177/0194599820950718.

[6] M.C. Martinez, A. Finegersh, F.M. Baik, F.C. Holsinger, H.M. Starmer, L.A. Orloff, J.B. Sunwoo, D. Sirjani, V. Divi, M.M. Chen, Comorbid Depression in Patients With Head and Neck Cancer Compared With Other Cancers, JAMA Otolaryngol. Neck Surg. 150 (2024) 1097–1104. 10.1001/jamaoto.2024.3233.

[7] P. Jimenez-Labaig, C. Aymerich, I. Braña, A. Rullan, J. Cacicedo, M.Á. González-Torres, K.J. Harrington, A. Catalan, A comprehensive examination of mental health in patients with head and neck cancer: Systematic review and meta-analysis, JNCI Cancer Spectr. (2024) pkae031. 10.1093/jncics/pkae031.

[8] A.M.H. Krebber, L.M. Buffart, G. Kleijn, I.C. Riepma, R. de Bree, C.R. Leemans, A. Becker, J. Brug, A. van Straten, P. Cuijpers, I.M. Verdonck-de Leeuw, Prevalence of depression in cancer patients: a meta-analysis of diagnostic interviews and self-report instruments, Psychooncology. 23 (2014) 121–130. 10.1002/pon.3409.

[9] S. Van Der Elst, Y. Bardash, M. Wotman, D. Kraus, T. Tham, The prognostic impact of depression or depressive symptoms on patients with head and neck cancer: A systematic review and meta-analysis, Head Neck 43 (2021) 3608–3617. 10.1002/hed.26868.

[10] X. Yang, G. Yang, R. Wang, Y. Wang, S. Zhang, J. Wang, C. Yu, Z. Ren, Brain glucose metabolism on [18F]-FDG PET/CT: a dynamic biomarker predicting depression and anxiety in cancer patients, Front. Oncol. 13 (2023) 1098943. 10.3389/fonc.2023.1098943.

[11] H. Da, N. Xiang, M. Qiu, S. Abbas, Q. Xiao, Y. Zhang, Characteristics of oxyhemoglobin during the verbal fluency task in subthreshold depression: A multi-channel near-infrared spectroscopy study, J. Affect. Disord. 356 (2024) 88–96. 10.1016/j.jad.2024.04.005.

[12] M.K. Yeung, J. Lin, Probing depression, schizophrenia, and other psychiatric disorders using fNIRS and the verbal fluency test: A systematic review and meta-analysis, J. Psychiatr. Res. 140 (2021) 416–435. 10.1016/j.jpsychires.2021.06.015.

[13] J.R. Binder, The Wernicke area, Neurology 85 (2015) 2170–2175. 10.1212/WNL.0000000000002219.

[14] G. Hickok, D. Poeppel, The cortical organization of speech processing, Nat. Rev. Neurosci. 8 (2007) 393–402. 10.1038/nrn2113.

[15] A.D. Craig, How do you feel — now? The anterior insula and human awareness, Nat. Rev. Neurosci. 10 (2009) 59–70. 10.1038/nrn2555.

[16] H.D. Critchley, S.N. Garfinkel, Interoception and emotion, Curr. Opin. Psychol. 17 (2017) 7–14. 10.1016/j.copsyc.2017.04.020.

[17] J.P. Hamilton, A. Etkin, D.J. Furman, M.G. Lemus, R.F. Johnson, I.H. Gotlib, Functional Neuroimaging of Major Depressive Disorder: A Meta-Analysis and New Integration of Baseline Activation and Neural Response Data, Am. J. Psychiatry (2012). 10.1176/appi.ajp.2012.11071105.

[18] T. Wise, J. Radua, E. Via, N. Cardoner, O. Abe, T.M. Adams, F. Amico, Y. Cheng, J.H. Cole, C. de Azevedo Marques Périco, D.P. Dickstein, T.F.D. Farrow, T. Frodl, G. Wagner, I.H. Gotlib, O. Gruber, B.J. Ham, D.E. Job, M.J. Kempton, M.J. Kim, P.C.M.P. Koolschijn, G.S. Malhi, D. Mataix-Cols, A.M. McIntosh, A.C. Nugent, J.T. O’Brien, S. Pezzoli, M.L. Phillips, P.S. Sachdev, G. Salvadore, S. Selvaraj, A.C. Stanfield, A.J. Thomas, M.J. van Tol, N.J.A. van der Wee, D.J. Veltman, A.H. Young, C.H. Fu, A.J. Cleare, D. Arnone, Common and distinct patterns of grey-matter volume alteration in major depression and bipolar disorder: evidence from voxel-based meta-analysis, Mol. Psychiatry 22 (2017) 1455–1463. 10.1038/mp.2016.72.

[19] M. Landgrebe, B. Langguth, K. Rosengarth, S. Braun, A. Koch, T. Kleinjung, A. May, D. de Ridder, G. Hajak, Structural brain changes in tinnitus: Grey matter decrease in auditory and non-auditory brain areas, NeuroImage 46 (2009) 213–218. 10.1016/j.neuroimage.2009.01.069.

[20] E. Vachon-Presseau, P. Tétreault, B. Petre, L. Huang, S.E. Berger, S. Torbey, A.T. Baria, A.R. Mansour, J.A. Hashmi, J.W. Griffith, E. Comasco, T.J. Schnitzer, M.N. Baliki, A.V. Apkarian, Corticolimbic anatomical characteristics predetermine risk for chronic pain, Brain 139 (2016) 1958–1970. 10.1093/brain/aww100.

[21] K.N. Fountoulakis, A. lacovides, S. Samolis, S. Kleanthous, S.G. Kaprinis, G.S. Kaprinis, P. Bech, Reliability, validity and psychometric properties of the Greek translation of the zung depression rating scale, BMC Psychiatry 1 (2001) 6. 10.1186/1471-244X-1-6.

[22] W.W.K. Zung, A Self-Rating Depression Scale, Arch. Gen. Psychiatry 12 (1965) 63–70. 10.1001/archpsyc.1965.01720310065008.

[23] M.E. Helidoni, T. Murry, J. Moschandreas, C. Lionis, A. Printza, G.A. Velegrakis, Cross-cultural adaptation and validation of the voice handicap index into Greek, J. Voice Off. J. Voice Found. 24 (2010) 221–227. 10.1016/j.jvoice.2008.06.005.

[24] L.R. Derogatis, K.L. Savitz, The SCL-90-R, Brief Symptom Inventory, and Matching Clinical Rating Scales, in: Use Psychol. Test. Treat. Plan. Outcomes Assess. 2nd Ed, Lawrence Erlbaum Associates Publishers, Mahwah, NJ, US, 1999: pp. 679–724.

[25] S. Donias, A. Karastergiou, N. Manos, Standardization of the symptom checklist-90-R rating scale in a Greek population, Psychiatriki 2 (1991) 42–48.

[26] G.E. Gignac, E.T. Szodorai, Effect size guidelines for individual differences researchers, Personal. Individ. Differ. 102 (2016) 74–78. 10.1016/j.paid.2016.06.069.

[27] W. Cheng, E.T. Rolls, J. Qiu, W. Liu, Y. Tang, C.-C. Huang, X. Wang, J. Zhang, W. Lin, L. Zheng, J. Pu, S.-J. Tsai, A.C. Yang, C.-P. Lin, F. Wang, P. Xie, J. Feng, Medial reward and lateral non-reward orbitofrontal cortex circuits change in opposite directions in depression, Brain J. Neurol. 139 (2016) 3296–3309. 10.1093/brain/aww255.

[28] W.C. Drevets, J.L. Price, M.L. Furey, Brain structural and functional abnormalities in mood disorders: implications for neurocircuitry models of depression, Brain Struct. Funct. 213 (2008) 93–118. 10.1007/s00429-008-0189-x.

[29] C.-T. Li, T.-P. Su, S.-J. Wang, P.-C. Tu, J.-C. Hsieh, Prefrontal glucose metabolism in medication-resistant majordepression, Br. J. Psychiatry 206 (2015) 316–323. 10.1192/bjp.bp.113.140434.

[30] S.M. Palmer, S.G. Crewther, L.M. Carey, The START Project Team, A Meta-Analysis of Changes in Brain Activity in Clinical Depression, Front. Hum. Neurosci. 8 (2015). 10.3389/fnhum.2014.01045.

[31] L. Su, Y. Cai, Y. Xu, A. Dutt, S. Shi, E. Bramon, Cerebral metabolism in major depressive disorder: a voxel-based meta-analysis of positron emission tomography studies, BMC Psychiatry 14 (2014) 321. 10.1186/s12888-014-0321-9.

[32] D. Bennabi, P. Vandel, C. Papaxanthis, T. Pozzo, E. Haffen, Psychomotor Retardation in Depression: A Systematic Review of Diagnostic, Pathophysiologic, and Therapeutic Implications, BioMed Res. Int. 2013 (2013) 158746. 10.1155/2013/158746.

[33] C.S.H. Ho, J. Wang, G.W.N. Tay, R. Ho, S.F. Husain, S.K. Chiang, H. Lin, X. Cheng, Z. Li, N. Chen, Interpretable deep learning model for major depressive disorder assessment based on functional near-infrared spectroscopy, Asian J. Psychiatry 92 (2024) 103901. 10.1016/j.ajp.2023.103901.

[34] L. Mao, X. Hong, M. Hu, Identifying neuroimaging biomarkers in major depressive disorder using machine learning algorithms and functional near-infrared spectroscopy (fNIRS) during verbal fluency task, J. Affect. Disord. 365 (2024) 9–20. 10.1016/j.jad.2024.08.082.

[35] R.G. Robinson, R.E. Jorge, Post-Stroke Depression: A Review, Am. J. Psychiatry (2016). 10.1176/appi.ajp.2015.15030363.

[36] C.L. McGrath, M.E. Kelley, P.E. Holtzheimer III, B.W. Dunlop, W.E. Craighead, A.R. Franco, R.C. Craddock, H.S. Mayberg, Toward a Neuroimaging Treatment Selection Biomarker for Major Depressive Disorder, JAMA Psychiatry 70 (2013) 821–829. 10.1001/jamapsychiatry.2013.143.

[37] J.A. Avery, W.C. Drevets, S.E. Moseman, J. Bodurka, J.C. Barcalow, W.K. Simmons, Major Depressive Disorder Is Associated With Abnormal Interoceptive Activity and Functional Connectivity in the Insula, Biol. Psychiatry 76 (2014) 258–266. 10.1016/j.biopsych.2013.11.027.

[38] H.S. Mayberg, Limbic-cortical dysregulation: a proposed model of depression, J. Neuropsychiatry Clin. Neurosci. 9 (1997) 471–481. 10.1176/jnp.9.3.471.

[39] M.D. Sidney H. Kennedy, M.S. Jakub Z. Konarski, P.D. Zindel V. Segal, P.D. Mark A. Lau, P.D. Peter J. Bieling, M.D. Roger S. McIntyre, M.D. Helen S. Mayberg, Differences in Brain Glucose Metabolism Between Responders to CBT and Venlafaxine in a 16-Week Randomized Controlled Trial, Am. J. Psychiatry (2007). 10.1176/ajp.2007.164.5.778.

[40] G. Hartwigsen, D. Saur, Neuroimaging of stroke recovery from aphasia – Insights into plasticity of the human language network, NeuroImage 190 (2019) 14–31. 10.1016/j.neuroimage.2017.11.056.

[41] J. Cole, S.G. Costafreda, P. McGuffin, C.H.Y. Fu, Hippocampal atrophy in first episode depression: A meta-analysis of magnetic resonance imaging studies, J. Affect. Disord. 134 (2011) 483–487. 10.1016/j.jad.2011.05.057.

[42] P. Videbech, B. Ravnkilde, Hippocampal Volume and Depression: A Meta-Analysis of MRI Studies, Am. J. Psychiatry (2004). 10.1176/appi.ajp.161.11.1957.

[43] C. Anacker, R. Hen, Adult hippocampal neurogenesis and cognitive flexibility — linking memory and mood, Nat. Rev. Neurosci. 18 (2017) 335–346. 10.1038/nrn.2017.45.

[44] E.J. Kim, B. Pellman, J.J. Kim, Stress effects on the hippocampus: a critical review, Learn. Mem. 22 (2015) 411–416. 10.1101/lm.037291.114.

[45] B.S. McEwen, P.J. Gianaros, Stress- and Allostasis-Induced Brain Plasticity, Annu. Rev. Med. 62 (2011) 431–445. 10.1146/annurev-med-052209-100430.

[46] C. Pittenger, R.S. Duman, Stress, Depression, and Neuroplasticity: A Convergence of Mechanisms, Neuropsychopharmacology 33 (2008) 88–109. 10.1038/sj.npp.1301574.

[47] M.A. Conway, C.W. Pleydell-Pearce, The construction of autobiographical memories in the self-memory system, Psychol. Rev. 107 (2000) 261–288. 10.1037/0033-295X.107.2.261.

[48] S.C. Prebble, D.R. Addis, L.J. Tippett, Autobiographical memory and sense of self, Psychol. Bull. 139 (2013) 815–840. 10.1037/a0030146.

